# Assessing Seroprevalence of SARS-CoV-2 Antibodies Among US College Students after School Reopening

**DOI:** 10.1101/2024.04.23.24305972

**Authors:** Scott E. Feeder, Tamar Ratishvili, Beth L. Carlson, Diane E. Grill, Krista M. Goergen, Richard B. Kennedy, Inna G. Ovsyannikova, Gregory A. Poland

## Abstract

**Objective:** This study assessed the seroprevalence of SARS-CoV-2 antibodies at two timepoints to evaluate the effectiveness of COVID-19 precautions in two colleges.

**Participants:** The study enrolled students, faculty, and staff from Covenant and Geneva Colleges.

**Methods:** During the fall of 2020 the Mayo Clinic Vaccine Research Group partnered with Vibrant America to hand out self-administered finger stick antibody tests at the start of the fall semester and again two months later.

**Results:** The study enrolled 305 participants from Covenant and 671 from Geneva College at timepoint 1 and 198 participants from Covenant and 554 from Geneva College at timepoint 2. Seroprevalence rates were 2.3% and 1.6% at timepoint 1, and 0.5% and 3.97% at timepoint 2 at Covenant and Geneva Colleges, respectively.

**Conclusion:** The results of this study suggest that implementation of strict preventive measures may have kept the spread of COVID-19 low at these colleges during the study period.

## INTRODUCTION

In March of 2020 the World Health Organization declared COVID-19 to be a global pandemic.^1^ In the same month the United States declared a National Public Health Emergency due to COVID-19 outbreaks.^2^ As COVID-19 infection rates continued to climb across the United States, several nonpharmaceutical interventions (NPIs) such as shelter-in-place orders, social distancing, mask wearing, and travel restrictions, were enacted including the closures of schools.^3^ Between March and April of 2020 the majority of schools had closed across all 50 states including K-12 schools, universities, and colleges affecting the spring semester.^4^ As the Fall school 2020 semester approached the United States President recommended that schools reopen for in-person learning,^5^ and national guidelines for reopening were put forward by the Centers for Disease Control and Prevention (CDC) and other professional organizations such as the American College Health Association.^6, 7^ The Chronicle of Higher Education reported that over a third of colleges and universities decided to allow students to return to campus in the fall of 2020, each implementing preventive measures and monitoring strategies of varying degrees of stringency.^8, 9^

Because of different factors including dense housing, large social gatherings, classroom and virtual learning, sporting events and other group activities, as well as varying degrees of concern and risk perception among college students and their parents, institutions of higher education (IHE), especially those with high-density residential housing were considered high-risk settings for the spread of infectious agents including SARS-CoV-2.^10–13^ In order to keep students, faculty, and staff safe during reopening, IHE which chose to resume on-campus learning had to plan for and implement policies and safety measures in accordance with federal, state, local, tribal, institutional, and other relevant health authority regulations.^14^ Considering the individual context of a college or university and consequently the need and practicality of specific COVID-19 prevention strategies, safety measures ranging from social distancing and mandatory masking to providing students, faculty, and staff with COVID-19 educational materials on a variety of screening and testing approaches were instituted.^15^ Notably, during the Fall of 2020 a unified guidance on SARS-CoV-2 testing in educational institutions was lacking and schools had to plan their strategies individually.

Several outbreaks were reported soon after the start of the Fall 2020 semester at a number of IHEs in the U.S. as well as worldwide.^16, 17^ These outbreaks provided opportunities to understand the true impact of IHE reopening and the relative effectiveness of preventive efforts against SARS-CoV-2 infection among students, faculty, and staff. However, case numbers and the magnitude of outbreaks can be underestimated in the absence of aggressive mass screening at frequent intervals, and these undetected cases can significantly contribute to disease spread in communities and limit outbreak control options.^18^ This is especially true for outbreaks at schools and IHE, as children, adolescents and young adults are more likely to have asymptomatic or minimally symptomatic SARS-CoV-2 infection and thus COVID-19 may go undetected in these age groups.^19, 20^ Therefore, an assessment of the true prevalence of SARS-CoV-2 among college/university students after resuming on-campus learning is crucial for informing the optimal strategies to minimize COVID-19 risks related to IHE.

Previous reports concerning COVID-19 prevalence in educational settings have mainly focused on detecting cases through PCR-based testing of symptomatic individuals and their contacts, generally targeted K-12 schools, involved single timepoint studies of seroprevalence and/or did not specifically address reopening.^21–29^ Our study aimed to identify trends in antibody seroprevalence in residential college students, faculty, and staff at two timepoints; at the beginning of the Fall 2020 semester and 2 months thereafter, to evaluate the prevalence and incidence of SARS-CoV-2 antibody positivity and the effectiveness of COVID-19 preventive measures (i.e., social distancing, mask wearing) put in place by campuses during the fall semester. Thus, the goal of this study was to assess baseline prevalence and incidence rates of SARS-CoV-2-specific antibodies among students, faculty, and staff approximately 8 weeks after reopening for in-person learning.

## METHODS

### Study Design and Subject Recruitment

During the summer of 2020 the Mayo Clinic Vaccine Research Group (VRG) based in Rochester, MN partnered with Vibrant America (San Carlos, CA) to design a study to ascertain SARS-CoV-2 antibody rates in college students, faculty, and staff returning to campus for the Fall 2020 semester. The VRG partnered with two colleges for this study: Covenant College located on Lookout Mountain in Dade County, GA and Geneva College located in Beaver Falls, PA. While both schools draw a small number of students from across the world and nationally, their primary demographic is regional.

Covenant College had an enrollment rate of 837 undergraduate students and 304 faculty and staff members on campus during the Fall of 2020. The college had 5 dormitories, including apartments, which housed 78% of the enrolled students. Since Covenant did not have separate IDs to distinguish between students and faculty/staff members; participants born before 1998 were categorized as faculty/staff and those born after 1998 as students (see Table 1 for demographic characteristics). If participants did not provide a student ID or date of brith, they could not be classified as a student or faculty/staff and where thus excluded from the study.

**Table 1:**
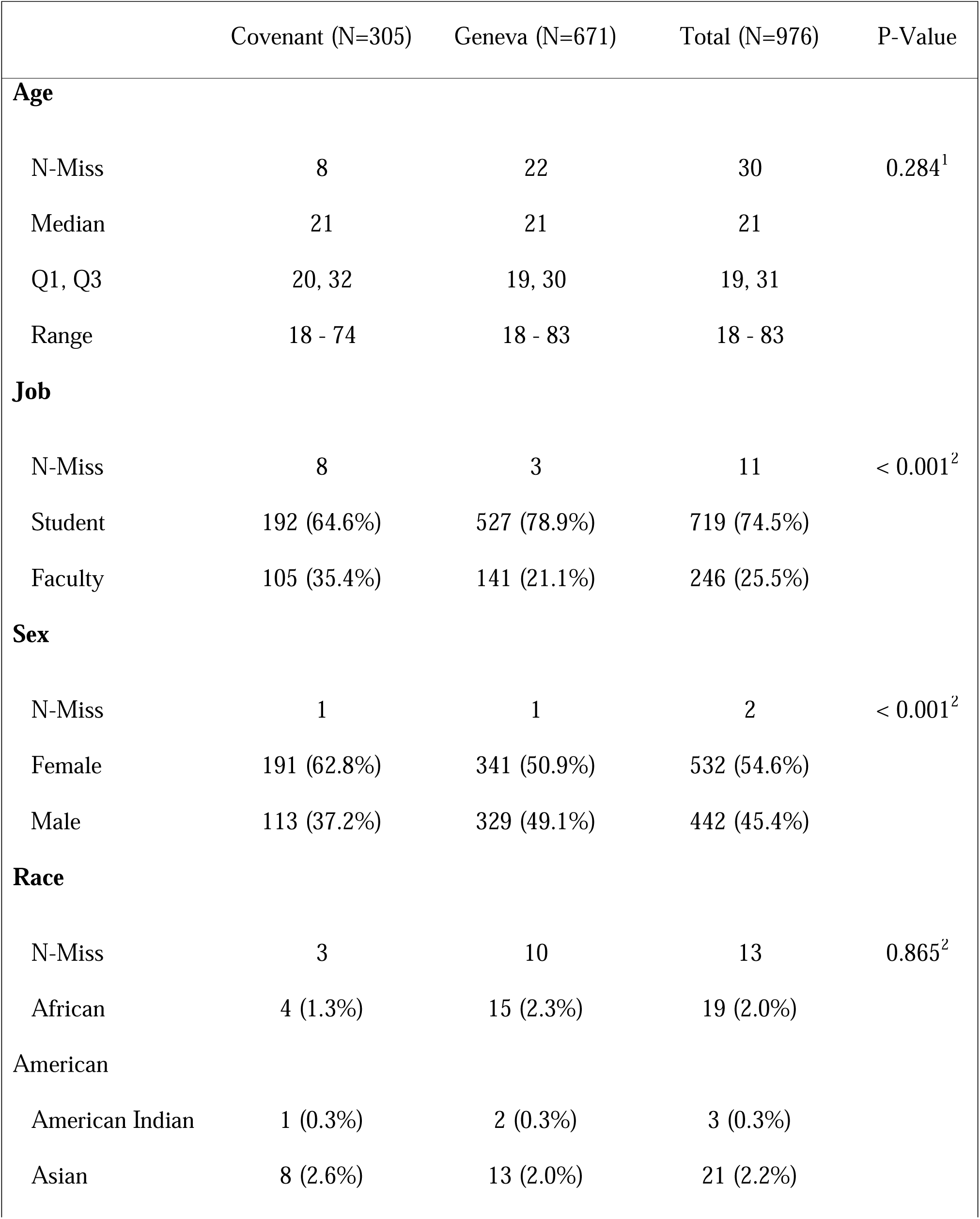
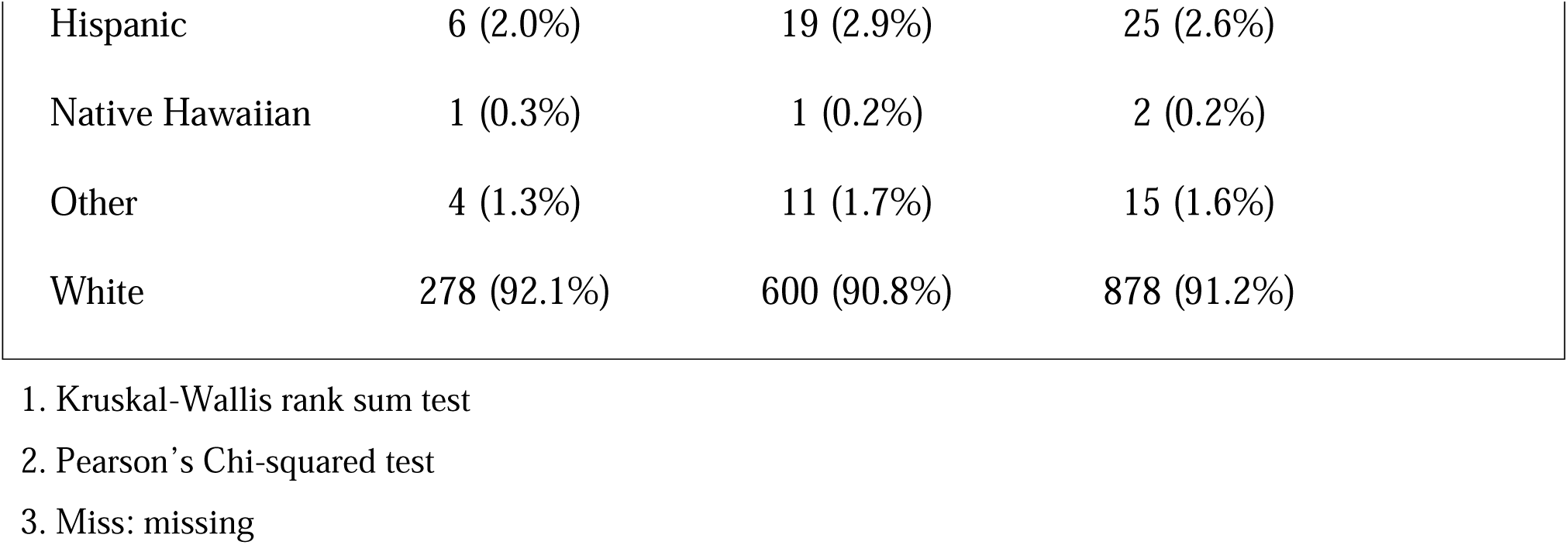
Demographic Characteristics of the Study Participants.

Geneva College had an enrollment rate of 965 undergraduate and 116 graduate students and 405 faculty and staff members on campus during the Fall of 2020. The campus had 9 residence halls which housed 73% of the enrolled students. Students and faculty/staff members were distinguished by college ID.

Testing for both colleges was completed at two timepoints. The first wave of testing started at the end of August 2020 as students, faculty, and staff returned to campus and ended in the first week of September 2020. A second wave of testing started during mid-October 2020 and ended mid-November 2020 (prior to Thangsgiving school breaks) and only included participants who completed the first wave of testing.

IRB approval was obtained from Mayo Clinic Rochester. Once approved, each College had their own IRB review and approve the study prior to recruitment. During the time of finger stick test kit distribution, participants were given a waiver of consent which explained the purpose of the study, how long they would be in the study (two testing periods), how data would be collected, stored, and used, any direct benefits to participant, and contact information for questions and concerns. No remuneration was offered with this study.

Vibrant America provided the finger stick kits which included an instruction booklet for completing the finger stick test. Shipping containers were also provided for sending the kits back to Vibrant America for testing. Participants either completed the testing themselves or could ask for assistance from a healthcare worker on campus. An online app was used to capture de-identified demographic information and conduct a brief questionnaire where study participants could self-report COVID-19 symptoms, any previous COVID-19 testing test type (nasal/PCR or serum antibody) and outcome (positive/negative). This information was paired with the antibody testing results during the analysis. The app was also used to provide participants the results of the Vibrant fingerstick antibody test and explanation of results.

Since each college campus is setup and run differently, a brief description of how each college implemented the study can be found in Appendix 1.

### SARS-CoV-2 Antibody (Ab) Testing

SARS-CoV-2 antibody assays were conducted on dried blood spot (DBS) samples self-collected by participants using finger stick kits. Antibody testing was completed on a Whatman 903TM DBS finger stick card (GE Healthcare Ltd. Chicago, IL) eluates using the Vibrant COVID-19 Ab chemiluminescence immunoassay (CLIA) developed for qualitative detection and differentiation of IgG and IgM antibodies directed at four major SARS-CoV-2 viral antigens (Wuhan strain): spike (S) [S1 and S2], receptor binding domain (RBD) and nucleocapsid (N) antigens. The Vibrant COVID-19 Ab Assay was granted an Emergency Use Authorization (EUA) by the FDA in June, 2020.^30, 31^ Test results are classified as positive or negative based on the reactivity of each sample calculated as the ratio of the sample intensity to the cut-off control intensity established by the manufacturer, with calculated values ≤ 1.0 indicating the absence of antibodies against SARS-CoV-2 (negative result) whereas values > 1.0 are considered preliminary for the presence of antibodies to one or more of the included antigens (positive result). The assay performed on serum specimens collected 4-26 days (median = 14 days) after a positive nasopharyngeal swab test demonstrated 98.11% (95% CI 90.06% - 99.67%) Positive Percent Agreement (PPA) with RT-PCR results. The Negative Percent Agreement (NPA) with SARS-CoV-2 antibody-negative serum specimens collected before the pandemic was reported as 98.60% (95% CI 97.14% - 99.32%). The total Coefficient of Variation (CV) for precision as well as lot to lot reproducibility was < 15%.

## RESULTS

Our study enrolled 305 (45%) participants from Covenant College and 671 (37%) from Geneva College at timepoint 1 from August 17 to September 16, 2020. The Covenant College cohort consisted of 35% faculty/staff members and 65% students. The Geneva College cohort consisted of 21% faculty/staff and 79% students.

For the second timepoint (visit 2) we enrolled 198 (26%) participants from Covenant College and 554 (17%) from Geneva College in the period from October 9 to November 17, 2020. Since participants needed to complete both timepoints 1 and 2 to be in the study, participants who only tested during time point 2 but had not completed the timepoint 1 visit were excluded.

As the COVID-19 CLIA assay measures antibodies to N, S1, S2 and RBD domains of SARS-CoV-2, SARS-CoV-2, IgM and IgG seropositivity were defined in our study as antibody positivity (value >1.00) to any one of the viral proteins. Table 2 below the results for IgG, IgM, self-reported PCR test, and previous antibody test results for timepoint 1.

**Table 2.**
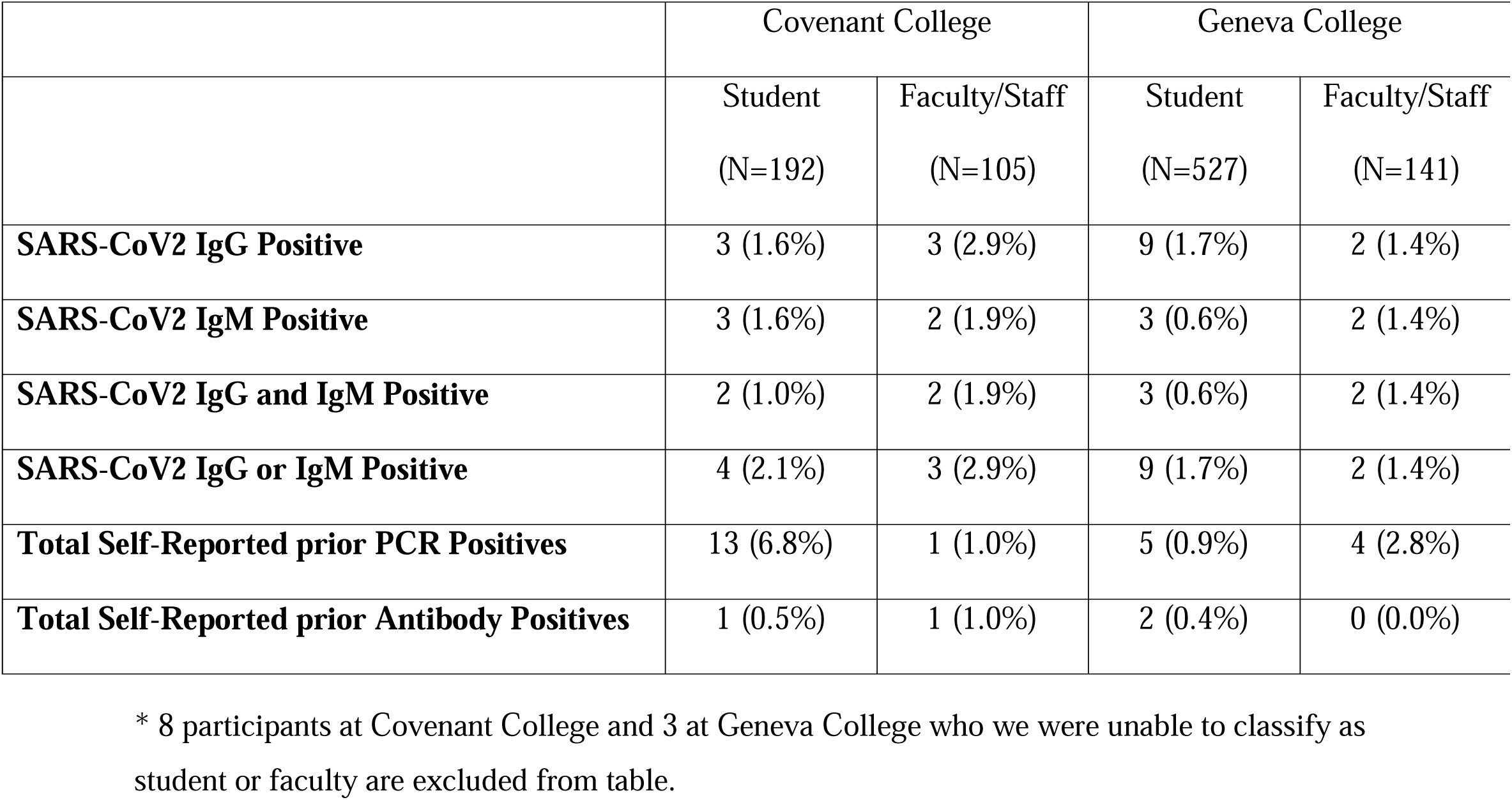
Antibody seropositivity, prior COVID-19 and antibody testing history at timepoint 1.

Overall antibody positivity at Covenant College at timepoint 1 was 2.3%. Of the 7 seropositive participants, 5 reported a positive PCR test and 1 reported a positive antibody test. At Geneva College, antibody positivity at visit 1 was 1.6%. Of the 11 seropositive participants, 3 reported a positive PCR test and 1 reported a positive antibody test.

Interestingly, of the 23 total individuals at both Colleges who self-reported PCR positivity, only 8 (34.8%) were seropositive (IgM or IgG) for at least one of the SARS-CoV-2 protein at timepoint 1.

With several exceptions, most IgM positive participants were positive for more than one SARS-CoV-2 antigen, with anti-RBD IgM antibodies being most prevalent with median values of 1.92 (IQR 1.36; 3.05) and 2.15 (IQR 1.47; 2.42) chemiluminescence units (CU) for Covenant and Geneva cohorts, respectively. Of the total of 10 IgM positive participants at both Colleges, 9 (90%) were positive for RBD-specific antibodies, while only 4 (40%) possessed N-specific antibodies. Notably, all 17 IgG seropositive participants demonstrated positive antibody responses to all four SARS-CoV-2 antigens included in the DBS testing assay. Table 3 shows the results for IgG, IgM, self-reported PCR test and previous antibody test results for timepoint 2.

**Table 3.**
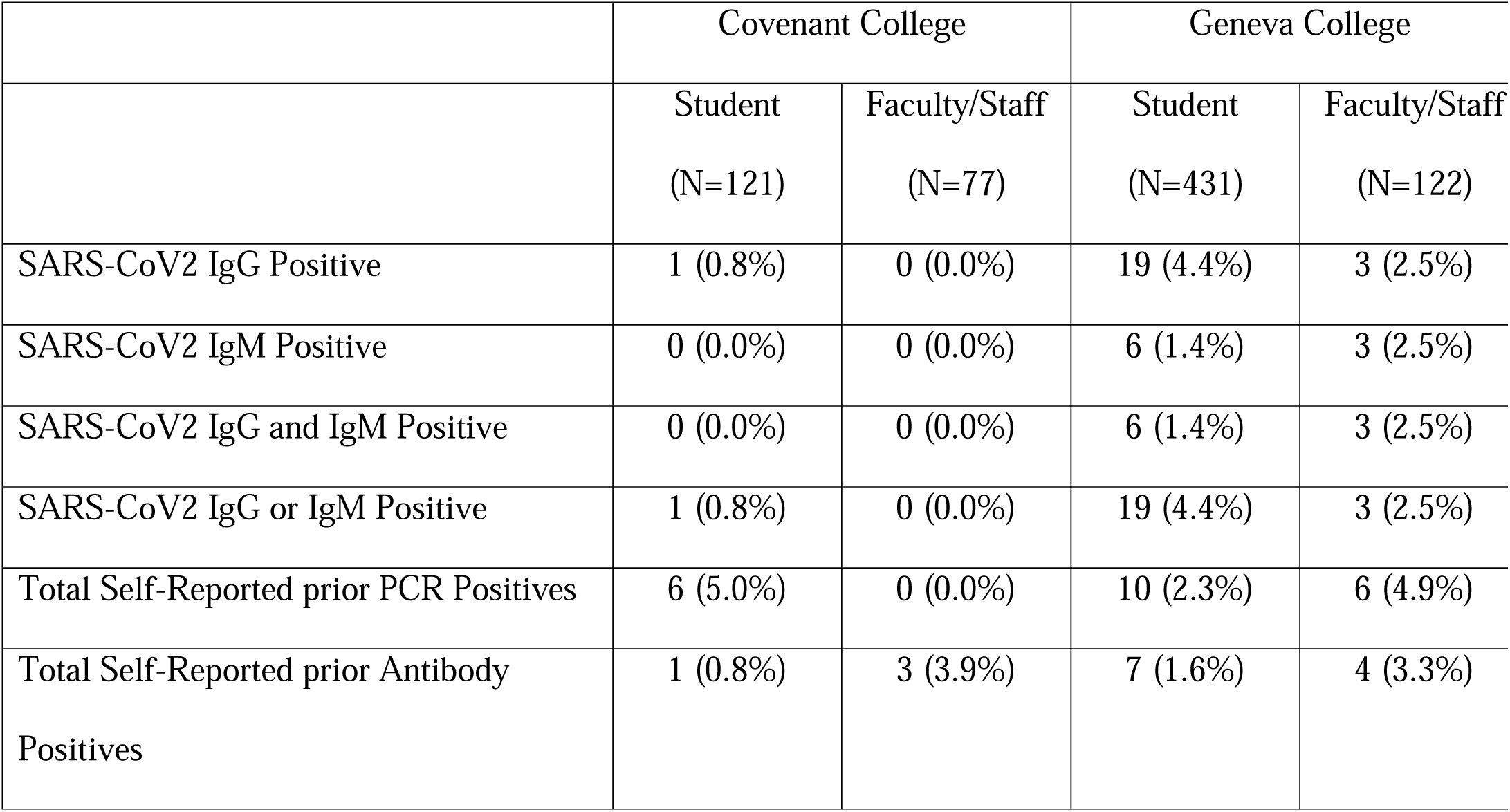
Antibody seropositivity, prior COVID-19 and antibody testing history at timepoint 2.

Only 1 participant from Covenant College tested positive for SARS-CoV-2 antibodies at time point 2. This individual also self-reported a positive PCR result. Notably, of the 7 participants identified as seropositive at timepoint 1, 3 did not participate in the second round of testing, while 4 participants were now seronegative.

Antibody positivity at Geneva College at timepoint 2 of the study was 3.97% with 16 new seropositive participants (i.e., participants who were SARS-CoV-2 antibody negative at time point 1). Of the total 22 seropositive participants, 11 (50%) reported a positive PCR test, and 4 (18.2%) reported a prior positive antibody test result. Of the 9 participants who were seropositive at timepoint 1 and returned for the second visit, 6 (66.7%) remained IgM or IgG seropositive.

Two previously seropositive participants did not return their second finger stick tests. Of all 22 subjects self-reporting a positive PCR test at both colleges by time point 2, only 12 (54%) were seropositive for SARS-CoV-2 antigens.

Similar to the trend observed at timepoint 1, 7 (77.8%) of 9 IgM positive participants were seropositive for RBD-specific antibodies, 4 (44.4%) for S1 and S2 domains of Spike protein, and only 3 (33.3%) were seropositive for N-specific IgM antibodies among participants at timepoint 2 from Geneva College. Of the 22 IgG positive participants from Geneva College, twenty (90.9%) were positive for IgG antibodies to all four included antigens, with 21 (95.5%) being seropositive for anti-S1 and 20 (90.9%) for anti-N, anti-S2 and anti-RBD. One seropositive participant at Covenant College was positive for IgG antibodies to N and S1 proteins at time point 2. Antibody response (in CUs) to one of any of the four antigens was positively correlated with antibody response to other antigens in both Geneva and Covenant College cohorts (Supplemental Figure 1).

Since our study was not able to enroll all students, faculty, and staff, and in order to assess the true magnitude of SARS-CoV-2 spread at colleges, at the conclusion of the study we requested data collected by the college health offices on the number of reported COVID-19 positives and those quarantined during the Fall semester (Table 4). The clinical cases documented by college nursing staff on the number of reported positive cases of COVID-19 reveals that our study did not capture the majority of these cases due to subject non-participation.

**Table 4.**
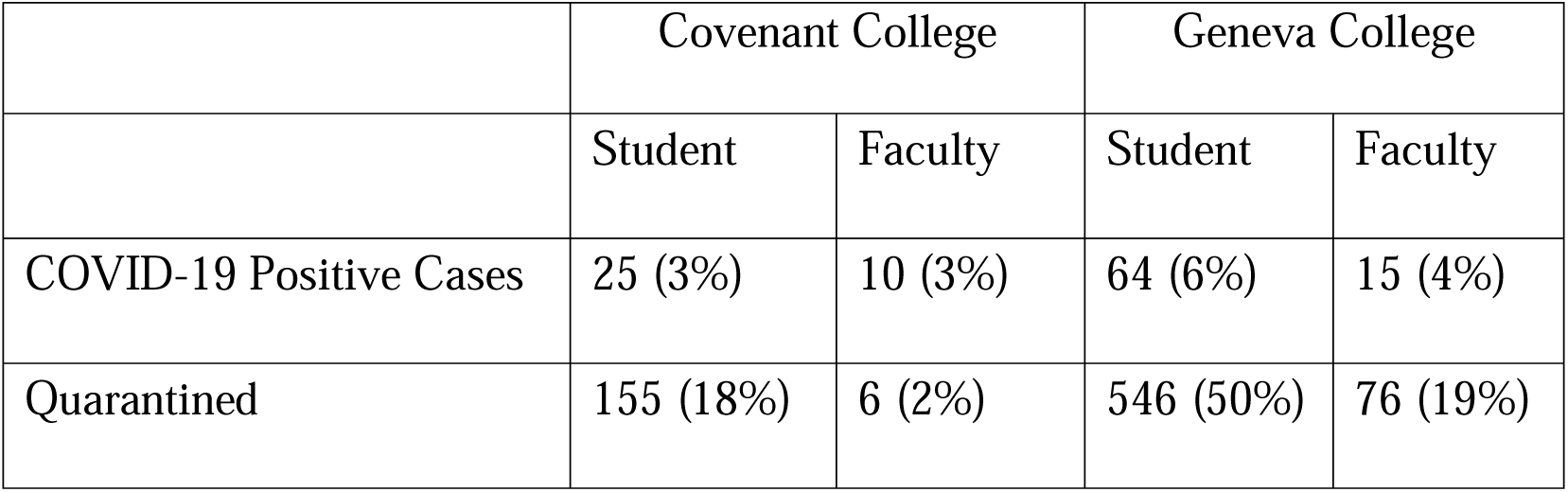
Total Students and Faculty with a Positive COVID-19 Test or Quarantined.

## DISCUSSION

Our two-timepoint study conducted among students, faculty, and staff members of two residential colleges from August to November 2020 assessed antibody seroprevalence to four major SARS-CoV-2 antigens using self-administered Dried Blood Spot (DBS) tests.

Unsurprisingly, from time point 1 to time point 2 we found an increase in the prevalence of seropositivity to SARS-CoV-2 antigens (as defined by antibody positivity to at least one of the antigens included in the DBS array [N, RBD, S1, S2]) of 1.6% to 3.97% among participants from Geneva College. This was likely related to the rise in COVID-19 incidence rates in Pennsylvania as well as most of the United States within the study period. This is also reflected in increased rates of self-reported positive PCR test results from 1.3% to 2.9%. However, in the Covenant College cohort measured antibody seropositivity and self-reported positive PCR test rates dropped from 2.3% to 0.8% and 4.6% to 3%, respectively within the study timeframe. This could, in part, be attributable to participants who tested positive for antibodies at visit 1 not returning for the 2^nd^ study time point. This decrease could also be linked to lower community rates of COVID-19 in the state of Georgia compared to Pennsylvania at the time and perhaps antibody waning (see below); however, our sample size is too limited to make this conclusion.

A considerable proportion of the seropositive individuals identified in our study did not report a previous positive SARS-CoV-2 PCR test result (Covenant College: 2/7 (28.5%) and 1/1 (100%); Geneva College: 8/11 (72.7%) and 11/22 (50%) at time points 1 and 2, respectively).

This is likely due to high rates of asymptomatic infection which account for approximately a third of all COVID-19 cases based on large meta-analyses.^32, 33^ The likelihood of developing symptoms following SARS-CoV-2 infection increases with age, with younger individuals demonstrating asymptomatic infections at a higher rate, reaching 80% in some reports.^34, 35^

SARS-CoV-2 seropositivity in our study cohort at both timepoints was similar to or lower as compared to reported seroprevalence rates among U.S. students as well as the general population in the same time frame. This aligns with findings of other studies done at educational settings during circulation of earlier SARS-CoV-2 variants and could suggest that with adequate countermeasures in place, it is realistic to keep in-school virus transmission rates, including asymptomatic transmission levels, similar to that of the community, despite the increased risk factors for transmission that are present on most college campuses.^25, 27, 29, 36^ It is, nevertheless, important to note that as seroprevalence rates vary greatly by geographic region, race, living conditions, occupation, income, socioeconomic status, climate, and the assay used for antibody detection, and perhaps most importantly, by variant of concern; therefore such rates should always be interpreted cautiously depending on such variables.^37–40^

It is known that a majority of individuals with COVID-19 develop detectable IgM and IgG antibodies to at least one of the major SARS-CoV-2 antigens independent of disease severity in the acute period following infection.^41–47^ Almost all seroconverted individuals develop IgM or IgG antibodies within 20 days of infection, with IgM levels declining rapidly afterwards and IgG levels often peaking later but persisting for months after infection. While most infected individuals exhibit IgM seroconversion followed by IgG seroconversion, other patterns of seroconversion, such as: synchronous IgM and IgG or IgM preceding IgG, have been reported.^41, 48^ Interestingly, a significant proportion (34.8% at time point 1 and 54% at time point 2) of study participants who self-reported a positive nasal swab PCR result were seronegative for SARS-CoV-2 antigen-specific antibodies. This could be due to several possible factors such as interindividual variations in the development and durability of antibody responses, timing between disease onset and sample collection, and assay limitations as follows. Following acute infection, persistence of SARS-CoV-2 antibodies over the period of months is variable depending on demographic factors, clinical course, antigen of interest, and the assay used for quantification, with some reports demonstrating rapid antibody waning and others showing high durability of antibody responses.^49–56^ As the dates of self-reported PCR and clinical diagnoses in our study cohort are unknown to us, antibody waning can at least partly explain why COVID-19 recovered participants subsequently tested seronegative during our study. Seronegativity in all individuals who self-reported positive PCR at timepoint 2 only (assumed to have had the disease in the interval between two study time points) was unexpected. As seroconversion can take up to three weeks after disease onset, ^43, 45, 57^ it is possible that samples of at least some study participants self-reporting PCR positivity had been self-collected early in the disease course before the development of detectable IgM or IgG antibodies. Albeit uncommon, the likelihood of false-positive PCR results should also be considered.^58–60^ Although DBS tests have been shown to reliably detect SARS-CoV-2 antibodies and their utility for seroprevalence studies has been validated,^61–64^ assay limitations such as the occurrence of false negative results in individuals with low antibody titers suggests that confirmatory testing on serum/plasma samples should be considered.^64, 65^

IgM and IgG antibody kinetics to different SARS-CoV-2 antigens also vary considerably. The vast majority of seroconverted individuals develop measurable antibodies to S1, N, and RBD protein domains.^66–71^ In our study, IgG antibody seropositivity was substantially higher than IgM seropositivity to SARS-CoV-2 antigens, which is in line with other reports as IgM titers to all SARS-CoV-2 antigens rapidly decrease following the acute phase of the disease.^72, 73^ Notably, IgG seropositivity to any one of the tested antigens was highly correlated with IgG seropositivity to the other three SARS-CoV-2 antigens in our cohort. However, IgM seropositivity to RBD antigen was more commonly observed in our cohorts than IgM seropositivity to S1, S2, and N. Several groups have reported faster decline of N-specific antibodies and longer persistence of RBD-induced antibody responses.^74–78^ Higher seroconversion rates to S1 and RBD domains versus nucleocapsid (N) protein in individuals with milder disease have also been observed.^50^ Moreover, Spike-based assays have been reported to surpass N-based assays in sensitivity of IgM detection.^79^ These factors could be contributing to a lower prevalence IgM of antibodies directed against N protein in our study. However, more insights into differential decline of IgM antibodies to different SARS-CoV-2 antigens and its possible biological significance are needed.

Overall, the increase in SARS-CoV-2 seropositivity as well as the rise in self-reported PCR positivity from time point 1 to time point 2 was unsurprising as COVID-19 cases continued to climb across the country and in both Georgia and Pennsylvania. According to the reported number of positive COVID-19 cases tracked by each of the College’s health services (Table 4), the true number of positive cases were higher than what was captured by this study (Tables 2 and 3). This is most likely explained by the lower-than-expected enrollment of students, faculty, and staff (Geneva 45% and Covenant 26%) at timepoint 1 and even lower retention numbers (Geneva 37% and Covenant 17%) at timepoint 2. Based on our seropositivity results and published literature on differences between observed and estimated actual numbers of COVID-19 cases in the United States, taking asymptomatic, minimally symptomatic, and untested individuals into account, we suggest that SARS-CoV-2 infection rates in both colleges were much higher than accounted for in our study or even the campus-wide numbers reported in Table 4.

The numbers of positive COVID-19 PCR tests provided by colleges reveal that more positive cases were reported at Geneva College as compared to Covenant College as outlined in Table 4. This makes sense for several reasons. First, Geneva had a larger enrollment of students and more faculty and staff members. Second, the possibility for exposure may have been greater at Geneva College as they have a graduate program. Graduate students may have lived off campus during the semester as compared to undergraduates and may have had more opportunities for exposure. Third, with a larger number of faculty and staff, Geneva College may have been at an increased risk of more off-campus exposure as faculty and staff were not living on campus. Importantly, there could have been differences in adherence to NPIs such as masking and social distancing between colleges.

Based on several reports to date, provided vigorous preventive measures are in place, educational institutions and dormitories usually do not represent major drivers of COVID-19 spread in communities.^80^ It has been shown that schools with multi-layered precautions and/or mask mandates have been more successful at limiting SARS-CoV-2 spread in classes and/or campuses and limiting outbreaks.^81–83^ Nevertheless, even though observations have been somewhat reassuring regarding a similar level of transmission in schools as in the community, some demonstrating even lower rates in children enrolled in on-site classes,^24, 84, 85^ several reports point to increases in county-level COVID-19 incidence following outbreaks at colleges with in-person instructional formats,^86, 87^ perhaps also indicative of differences between K-12 schools and higher educational settings. Further, during the period of our study COVID-19 vaccines were not yet authorized. Safe and effective vaccines are now available to everyone 5 and older, including students and faculty of institutions of higher education, which greatly lowers the risks of large outbreaks at colleges and in communities with high vaccination coverage.^88^

The emergence of new SARS-CoV-2 variants of concern (VOCs) is another factor to consider in the context of COVID-19 infections in educational settings due to higher transmissibility of emerging variants compared to the original (Wuhan-Hu-1 or USA/WA1/2020) and earlier VOCs that predominated during the previous academic year. This remains important in immunized students as well due to lower susceptibility of emerging variants to vaccine- and illness-induced immunity, specifically, lower magnitude of neutralization by both infection and vaccine-induced antibodies, the effect most pronounced with Omicron variant.^89–91^ The problem of waning of vaccine-induced antibodies should also be factored in as protection conferred by vaccination can also decrease over time in immunized students, faculty, and staff members, highlighting the importance of continuation of non-pharmaceutical precautions.^92^

While two-timepoint seroprevalence studies have been conducted in K-12 schools worldwide and in non-U.S. colleges/universities, to the best of our knowledge, this is the first study of this kind conducted in U.S. colleges at the time of the Fall 2020 semester reopening.

The information obtained from our study regarding prevalence of SARS-CoV-2 antibodies at the start of the Fall 2020 semester and 2 months later will hopefully provide knowledge on viral spread in the face of various preventive measures in place on campuses and can help inform future decision-making and planning.

The strengths of our study included the fact that even though enrollment and retention were lower than expected (45% and 37% at time point 1; 26% and 17% at time point 2 for Covenant and Geneva Colleges, respectively), timepoint 1 did give us a sampling of SARS-COV-2 antibody titer results for a sizeable number of returning students, faculty, and staff. These results were presented to each college to help guide them, if desired, in policy and safety precautions on their campuses. It is unknown how the results were interpreted by each college and if COVID-19 policy or practices were changed or updated as a result. Our study also benefited from the use of a self-administered finger stick test. The test itself is less invasive than a venipuncture blood draw and can be completed by the participant at their leisure and from their preferred location, demonstrating the utility of such self-administered assays in future outbreaks.

This study has several limitations that should be considered and could be addressed in future studies of similar design. First, enrollment and retention rates in the study were lower than expected. This could partly be explained by the relative novelty of COVID-19 and attendant testing and safety precautions at the period. Another plausible explanation could be that the waiver of consent did not offer enough education on the benefit of knowing one’s SARS-CoV-2 antibody titer and thus participants were not interested in the study. Second, the fingerstick test itself may have led to low recruitment and retention. Some may have found the concept of self-pricking their finger not appealing or found that it was painful or difficult and declined a second testing. Third, participants may have been satisfied with their antibody result at timepoint 1 and had no interest in completing a second testing. Fourth, no remuneration was offered for this study which also could have affected enrollment and retention. Lastly, the recruitment effort was very limited on resources and availability as the new semester started. The nursing staff that led this effort had little extra time available while trying to prepare the campus for the return of students and faculty during the pandemic.

In addition, the results of the study are limited in that they represent two unique small Christian liberal arts colleges with lower student, faculty, and staffing numbers as compared to larger public state universities. Additionally, as stated above, the administered questionnaire did not include questions regarding dates of reported infection or PCR tests which would be beneficial in interpreting seroprevalence rates in these individuals. Another important factor not addressed in the questionnaire was the clinical course of the reported infection, which would aid us in comparing our results with other reports of antibody seroprevalence based on disease presentation.

## CONCLUSION

In conclusion, this study revealed increased rates of seropositivity to SARS-CoV-2 at study timepoint 2. Regardless, the occurrence of COVID-19 cases in institutions of higher education is inevitable. However, we believe that the implementation of strict preventive measures ranging from mandatory masking to quarantine requirements seem to be able to keep these risks close to that of the general population in the setting of high nationwide community transmission and surge of COVID-19 cases during the study period.

Currently, we are unaware of another research group attempting to track COVID-19 seroprevalence in US institutions of higher education during the start and end of a semester. Studies like these are important for better understanding whether current COVID-19 precautions are sufficient or should be reassessed. We hope that our approach and study design will be of interest and benefit for others studying the prevalence and spread of SARS-CoV-2 or other emerging pathogens in select populations.

## Supporting information

Manuscript Tables

## Data Availability

All data produced in the present study are available upon reasonable request to the authors

## Acknowledgements

Covenant College and Geneva College, health Staff: Peyton B Carlson, MPAS, PA-C, Rebecca Mack RN, BSN and student workers: Connor Gilchrest, Audrey Kenney, Jacob Neiderhiser, Katryn Gatchell, and also the students and faculty and the President and administrators of these schools. Vibrant America: Vasanth Jayaraman.

## Appendices

## Appendix 1. Study implementation for each college

### Geneva College

The study was advertised in the student and parent “FYI” news email sent out prior to arriving on campus in August 2020. Each student was also given a study flyer during the dorm check-in time. Faculty and staff also received study flyers. Students, Faculty, and staff were also reminded to complete the study several additional times with study flyers put in their campus mailboxes and via college emails. Information was also posted on Geneva’s health services social media accounts (Facebook and Instagram).

Finger stick kits and waivers of consent were distributed by health services staff and nursing students from a tent setup on the campus grounds. Participants were informed about a drop-box outside of the campus mail room to deposit test kits. Kits were then sent back to Vibrant America for testing.

During the second wave of testing, a distribution center was setup in a campus building or via tents, weather permitting. Qualifying participants were sent emails during the second wave of testing reminding participants to complete the finger stick test. Lastly, finger stick test kits were placed in campus mailboxes of those who still had not completed the study during the final days of testing.

### Covenant College

The study was advertised to students, faculty, and staff through email, campus study flyers, and an article printed in the student newspaper. Finger stick kits were picked up and dropped off at several locations around campus. Finger stick kits and waivers of consent were distributed by Student Development and Health Services staff.

During the second wave of testing, finger stick kits were picked up and dropped off at the same locations around campus. Qualifying participants were sent emails during the second wave of testing reminding participants to complete the finger stick test. Lastly, finger stick test kits were placed in campus mailboxes of those who still had not completed the study during the final days of testing.

Each college had their own approach to preparing and advocating for COVID-19 precautions during the Fall semester. These precautions fit into 6 criteria (screening, sanitization, social distancing, quarantining, masking, and college culture) which are outlined in Appendix 2.

## Appendix 2. Covenant College COVID-19 precautions

### COVID-19 Screening

Students needed to have a negative PCR nasal swab test or negative antigen test 5-7 days prior to their arrival date on campus. Students could also present a positive case from the previous 90 days (not to include the previous 10 days) or a positive antibody test from the past 30 days. Once on campus, all students, faculty, and staff were asked to review COVID-19 screening questions and take their temperature daily access for COVID-19 symptoms, daily.

### Sanitization

All students, faculty and staff were asked to follow public health practices including frequent handwashing or usage of alcohol-based hand sanitizer, proper respiratory etiquette including covering coughs and sneezes, and observing all rules for wiping down equipment and workstations on campus as directed.

### Social Distancing

All students, faculty and staff were asked to stay at least six feet apart from others and eliminate nonessential physical touch (for example, replace handshakes and hugs with elbow bumps). All classrooms were set up to permit six feet of distance between seats. Plexiglass shields were installed in high traffic/high concentration service areas to protect the most vulnerable.

### Quarantining

Students diagnosed who tested positive for COVID-19 were required to quarantine. Students who reported close contact with or cared for someone with documented or suspected COVID-19 within the last 14 days or had a new onset of symptoms consistent with viral illness such as fever of 100.4° or greater, cough, shortness of breath or difficulty breathing, chills, muscle pain, sore throat, nausea/diarrhea, or new loss of taste or smell were told to remain in their place of residence and monitor themselves for other symptoms. Along with empty rooms on each hall, the college set aside two halls, three apartments, and rental houses where students who tested positive or were symptomatic could be isolated should they be unable to return home.

### Masks

All students, faculty and staff were required to wear face coverings when indoors in public areas.

### College Culture

As a Christian College, Covenant communicated to students, faculty, and staff it was because they did not want to get others sick and that they should faithfully maintain good hygienic practices, daily self-screening, and stay home if necessary and it was out of concern for the most vulnerable that they wear masks and physically distance themselves when the situation called for it, regardless of personal preferences or convictions.

A document called the Covenant Commitment was created by Covenant College outlining the precautions above was handed out to students, staff, and employees. This information was also made available electronically on the college’s website.

At the conclusion of this study, campus staff acknowledged that students, staff, and employees overall adhered to the standards set forth. However, they acknowledged that these standards were not fully upheld by all members on campus.

*Geneva College COVID-19 precautions*

### COVID-19 Screening

All members of the Geneva College community, including students, faculty, staff, visitors, and independent contractors, were asked to review COVID-19 screening questions and take their temperature daily. Unlike Covenant College, Geneva did not require students to have a negative PCR or antibody SARS-CoV-2 test prior to returning to campus.

### Sanitization

Hand sanitizer stations were placed at entrances and exits to every building. Cleaning products were made available to wipe down tables and desks before and after classes.

### Social Distancing

All students, faculty, and staff were asked to stay at least six feet apart to avoid SARS-CoV-2 exposure and slow its spread. Plexiglass dividers were installed in cafeteria areas and seats were spaced at 6-foot intervals. A 10-12-foot buffer space was put in place between faculty to the first row of students in each classroom.

### Quarantining

If a student answered yes to a COVID-19 screening question (i.e., a fever ≥ 100.4° F) with either a temporal or oral thermometer, and/or symptoms of acute respiratory illness (i.e. dry cough, shortness of breath, new onset of lack of taste and/or smell, new onset of nasal congestion or runny nose, new onset of nausea, vomiting, diarrhea, chills or repeated shaking chills, headache, sore throat, and/or muscle aches) the student was quarantined in a quarantine room and roommates were quarantined in their house or apartment until the sick student’s test was reported. If the sick student was in a traditional dorm with a shared bathroom, then the roommate was also placed in a quarantine room with their own bathroom until the sick roommate’s COVID-19 test results were available. If the sick roommate tested positive, then all roommates and contacts were tested for COVID-19 via PCR testing.

### Masks

All students, staff, faculty, and independent contractors were asked to wear a mask while on campus parameters.

### College Culture

As a Christian College, Geneva communicated to students, faculty, and staff that they are committed to cultivating a living and learning community consistent with their mission to equip students for faithful and fruitful service to God and neighbor but also stressed the need to be vigilant, unified, and follow CDC safety guidelines.

A document outlining the precautions above was handed out to students, staff, and faculty. This information was also made available electronically on the college’s website. At the conclusion of this study, campus staff acknowledged screening efforts by students, staff, faculty as poor starting about mid-semester.

**Supplemental Figure 1.**
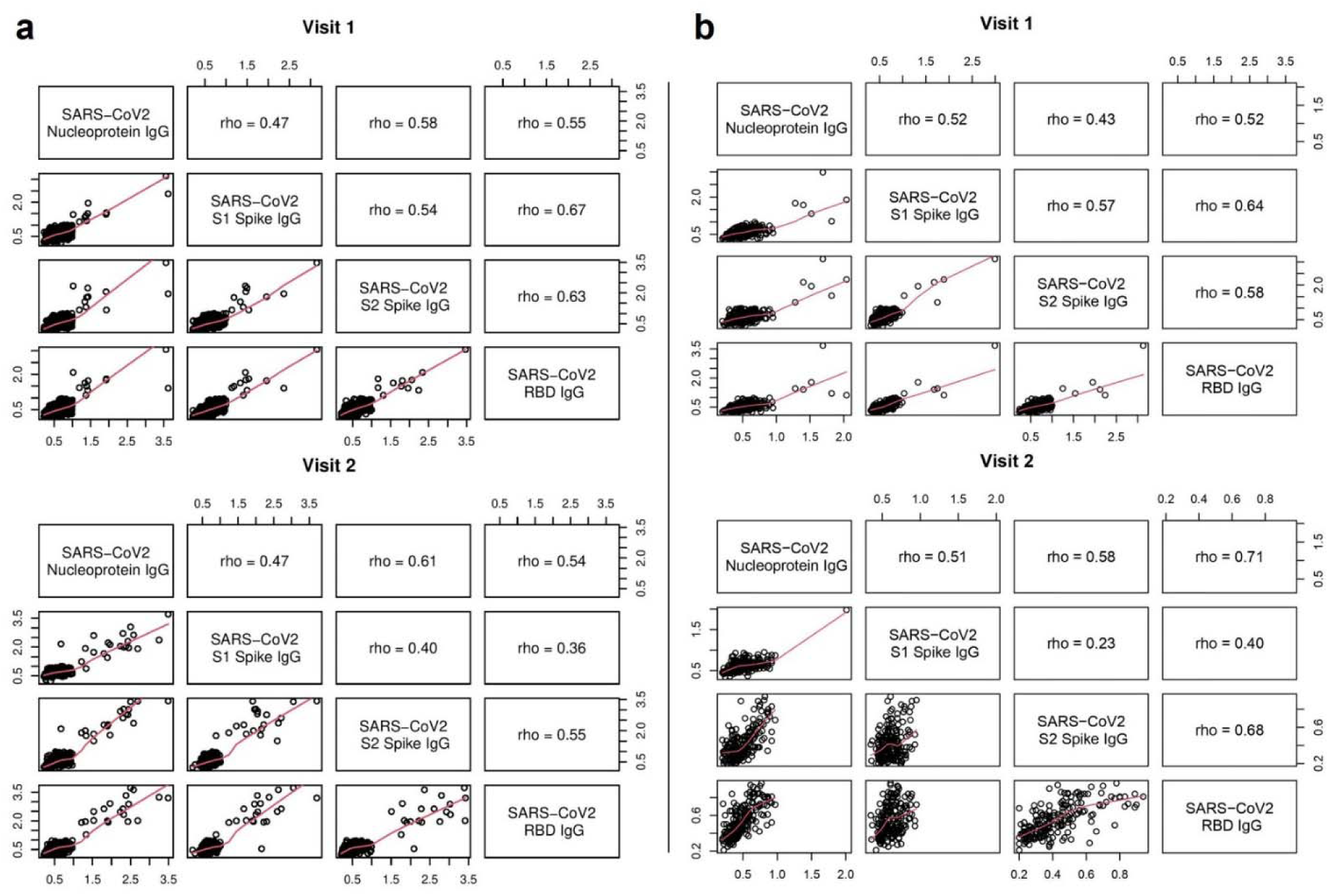
Correlations between antibody responses (in CUs) to each of the tested antigens in Geneva (**a**) and Covenant (**b**) cohorts at two timepoints

