## Supplementary material for "Assessing Seroprevalence of SARS-CoV-2 Antibodies Among US College Students after School Reopening": Manuscript Tables

|  | Covenant (N=305) | Geneva (N=671) | Total (N=976) | P-Value |
| --- | --- | --- | --- | --- |
| **Age** |  |  |  |  |
| N-Miss | 8 | 22 | 30 | 0.284^1^ |
| Median | 21 | 21 | 21 |  |
| Q1, Q3 | 20, 32 | 19, 30 | 19, 31 |  |
| Range | 18 - 74 | 18 - 83 | 18 - 83 |  |
| **Job** |  |  |  |  |
| N-Miss | 8 | 3 | 11 | < 0.001^2^ |
| Student | 192 (64.6%) | 527 (78.9%) | 719 (74.5%) |  |
| Faculty | 105 (35.4%) | 141 (21.1%) | 246 (25.5%) |  |
| **Sex** |  |  |  |  |
| N-Miss | 1 | 1 | 2 | < 0.001^2^ |
| Female | 191 (62.8%) | 341 (50.9%) | 532 (54.6%) |  |
| Male | 113 (37.2%) | 329 (49.1%) | 442 (45.4%) |  |
| **Race** |  |  |  |  |
| N-Miss | 3 | 10 | 13 | 0.865^2^ |
| African American | 4 (1.3%) | 15 (2.3%) | 19 (2.0%) |  |
| American Indian | 1 (0.3%) | 2 (0.3%) | 3 (0.3%) |  |
| Asian | 8 (2.6%) | 13 (2.0%) | 21 (2.2%) |  |
| Hispanic | 6 (2.0%) | 19 (2.9%) | 25 (2.6%) |  |
| Native Hawaiian | 1 (0.3%) | 1 (0.2%) | 2 (0.2%) |  |
| Other | 4 (1.3%) | 11 (1.7%) | 15 (1.6%) |  |
| White | 278 (92.1%) | 600 (90.8%) | 878 (91.2%) |  |

**Table 1:** Demographic Characteristics of the Study Participants

1. Kruskal-Wallis rank sum test

2. Pearson’s Chi-squared test

3. Miss: missing

|  | Covenant College | | Geneva College | |
| --- | --- | --- | --- | --- |
|  | Student (N=192) | Faculty/Staff (N=105) | Student (N=527) | Faculty/Staff (N=141) |
| **SARS-CoV2 IgG Positive** | 3 (1.6%) | 3 (2.9%) | 9 (1.7%) | 2 (1.4%) |
| **SARS-CoV2 IgM Positive** | 3 (1.6%) | 2 (1.9%) | 3 (0.6%) | 2 (1.4%) |
| **SARS-CoV2 IgG and IgM Positive** | 2 (1.0%) | 2 (1.9%) | 3 (0.6%) | 2 (1.4%) |
| **SARS-CoV2 IgG or IgM Positive** | 4 (2.1%) | 3 (2.9%) | 9 (1.7%) | 2 (1.4%) |
| **Total Self-Reported prior PCR Positives** | 13 (6.8%) | 1 (1.0%) | 5 (0.9%) | 4 (2.8%) |
| **Total Self-Reported prior Antibody Positives** | 1 (0.5%) | 1 (1.0%) | 2 (0.4%) | 0 (0.0%) |

**Table 2.** Timepoint 1

* 8 participants at Covenant College and 3 at Geneva College who we were unable to classify as student or faculty are excluded from table.

|  | Covenant College | | Geneva College | |
| --- | --- | --- | --- | --- |
|  | Student (N=121) | Faculty/Staff (N=77) | Student (N=431) | Faculty/Staff (N=122) |
| SARS-CoV2 IgG Positive | 1 (0.8%) | 0 (0.0%) | 19 (4.4%) | 3 (2.5%) |
| SARS-CoV2 IgM Positive | 0 (0.0%) | 0 (0.0%) | 6 (1.4%) | 3 (2.5%) |
| SARS-CoV2 IgG and IgM Positive | 0 (0.0%) | 0 (0.0%) | 6 (1.4%) | 3 (2.5%) |
| SARS-CoV2 IgG or IgM Positive | 1 (0.8%) | 0 (0.0%) | 19 (4.4%) | 3 (2.5%) |
| Total Self-Reported prior PCR Positives | 6 (5.0%) | 0 (0.0%) | 10 (2.3%) | 6 (4.9%) |
| Total Self-Reported prior Antibody Positives | 1 (0.8%) | 3 (3.9%) | 7 (1.6%) | 4 (3.3%) |

**Table 3.** Timepoint 2

**Table 4.** Total Students and Faculty with a Positive COVID-19 test or Quarantined

|  | Covenant College | | Geneva College | |
| --- | --- | --- | --- | --- |
|  | Student | Faculty | Student | Faculty |
| COVID-19 Positive Cases | 25 (3%) | 10 (3%) | 64 (6%) | 15 (4%) |
| Quarantined | 155 (18%) | 6 (2%) | 546 (50%) | 76 (19%) |
